# Assessment of validity and reliability of the Tuberculosis Related Stigma Scale in Colombian patients

**DOI:** 10.1101/2021.12.01.21267141

**Authors:** Neddy Pamela Castañeda-Daniels, Adalberto Campo-Arias, John Carlos Pedrozo-Pupo

## Abstract

**Objective:** To know the dimensionality and internal consistency of the Tuberculosis-Related Stigma Scale in patients living with tuberculosis in Santa Marta, Colombia. Method: One hundred and twenty-two patients between the ages of 18 and 75 participated (M=40.3, SD=14.9), 63.9% were men, 44.3% were single, 69.7% had low income, 80.3% had pulmonary tuberculosis, and 13.1% had co-infection with HIV. The Tuberculosis-Related Stigma Scale was applied; it is composed of two subscales: perceived and internalized stigma. The internal structure was explored by confirmatory factor analysis (EFA). Internal consistency was measured with Cronbach’s alpha and McDonald’s omega. Besides, the differential functioning of the scales according to gender was explored with Kendall’s tau-b coefficient.

**Results:** CFA did not show excellent goodness-of-fit indicators for the perceived stigma scale (Satorra-Bentler’s chi-square of 184.48, degree of freedom of 44, p=0.001, RMSEA of 0.16, 95%CI 0.14 - 0.19, CFI of 0.77, TLI of 0.72, and SRMR of 0.08) and internalized (Satorra-Bentler’s chi-square of 189.14, degree of freedom of 54, p=0.001; RMSEA of 0.14, 95%CI 0.12 - 0.17, CFI of 0.82, TLI of 0.78, and SRMR of 0.07). The alpha and omega coefficients were 0.89 and 0.91 for both scales, respectively. Non-gender differential functioning was observed; Kendall’s tau-b were between 0.00 and 0.15.

**Conclusions:** The Tuberculosis-Related Stigma Scale has an excellent internal consistency but poor goodness-of-fit indicators of unidimensionality. Evaluating the scale’s psychometric performance is recommenced in future research.

## Introduction

Tuberculosis (TB) is an infectious disease that usually affects primarily lung tissue and, secondarily, tissues of other organs^1^. TB is one of the leading causes of death globally; however, preventive measures and effective treatment since people develop TB can be successfully treated with a six-month treatment regimen^2^. It is noteworthy that ending tuberculosis by 2030 is part of the sustainable development goals implemented by the World Health Organization^3^.

In Colombia, strictly supervised shortened treatment was adopted to improve adherence to TB treatment^4^. However, success has been limited, given that an incidence of 18,000 cases per 100,000 inhabitants is still observed^5^. The social determinants of health are possibly the most significant barrier to controlling and eradicating the disease^6^. Among the access barriers for the control of endemic diseases such as TB are 1) Cost of medicines, medical consultations, and exams; 2) Fear or shame when being treated in health service; 3) distrust of health teams and prescribed treatment; and 4) stigma-discrimination, beliefs, and myths^7^. The “End TB Strategy” emphasizes the stigma-discrimination associated with TB and the role of stigma-discrimination in hindering efforts to combat TB^3^.

Stigmatization is conceptually different from discrimination, prejudice, and stereotype. Stigma is defined as the process by which a person is devalued or discredited based on an attribute that gives them an undesirable connotation^8^. Unlike prejudice, which refers to the individual mental process, the characteristic indicated as undesirable or with little value is accepted^9^. The stereotype is more related to the devalued image accepted and generalized to most societies^10^. Furthermore, discrimination is all the behaviours that imply the exclusion, marginalization, and violation of the rights of people who were stigmatized and about whom a prejudice persists^9^.

The stigma-discrimination complex related to TB (TB-SDC) is understood as the negative assessment that the person has of himself due to the experience of living with TB^10^. Internalized stigma is related to the individual’s internal perceptions and emotions^11^. Perceived stigma refers to the fear of discrimination or, in general, awareness of negative attitudes or practices related to a particular condition that makes it impossible for people to comment on their experiences and feelings of guilt and shame^12^. TB is stigmatized because it is linked to other devalued social characteristics^13^. TB is a syndemic associated with poverty, malnutrition, abuse of alcohol and psychoactive substances, incarceration, co-infection with the human immunodeficiency virus, and recent migration from countries with a high disease burden^5^.

The TB-SDC is one of the leading social factors that cause a delay in diagnosis and non-compliance with treatment among people who live with TB^14^. Additionally, TB-SDC increases the risk of depressive disorders^15^. A situation that further impairs treatment adherence^13^. In general, TB-SDC deteriorates patients’ quality of life since it leads to isolation, fewer opportunities to get and keep a job, and delays the search for medical attention^13^.

The growing importance of TB-SDC is reflected in instruments available for the quantification of the construct, such as the Scale of Stigma Related to Tuberculosis^16^, adapted for Portuguese in Brazil^17^, and Spanish in Mexico^18^, the Scale of Coreil Stigma in the United States^11^, and an instrument to measure stigma towards TB in Colombia^19^.

The Tuberculosis-Related Stigma Scale comprises 23 items, 11 of them to explore the community perspective towards TB (perceived stigma sub-scale) and the other of 12 items to know the patient’s vision (sub-scale internalized stigma). The scale offers a polytomous response pattern with four options, the higher the score, the higher the stigma. Two samples of 204 and 480 people who live with TB showed high internal consistency for both subscales, 0.88 and 0.82, respectively. However, dimensionality, 30-day stability, and nomological validity were lacking^16^.

In Mexico, a translation and adaptation of the Tuberculosis-Related Stigma Scale was carried out in 217 patients and observed high internal consistency for the subscales was satisfactory, Cronbach’s alpha of 0.88 for the perceived stigma subscale 0.91 for internalized stigma. The exploratory factor analysis, separated by subscales, showed poor dimensionality^18^.

Subsequently, the Tuberculosis Related Stigma Scale was translated into Brazilian Portuguese. In a sample of 83 people who live with TB, the perceived stigma subscales showed acceptable internal consistency, both with Cronbach’s alpha of 0.70, without informing the dimensionality analysis^17^. In Turkey, the Turkish version of the Tuberculosis-Related Stigma Scale showed that in 263 participants without a TB diagnosis, they observed that the perceived stigma sub-scale showed Cronbach’s alpha of 0.83 and internalized stigma 0.89. From the confirmatory factor analyzes, they showed acceptable values for dimensionality^18^.

In Colombia, the internal consistency and factor solution of the Tuberculosis-Related Stigma Scale subscales are unknown. The two subscales show Cronbach’s alpha greater than 0.70 in the available studies. However, in the factor analysis, the findings are not significant due to the poor performance of some items since it gives the impression that the factor analysis was carried out jointly for the two subscales^16-18^. Given the characteristics of the construct, it was advisable to do an independent factorial analysis for each subscale^20^.

A reliable instrument guarantees the investigations’ internal validity that determines the prevalence and other variables associated with TB-SDC.20 This more robust understanding of the role of the TB-SDC will help the design of actions aimed at reducing the perceived stigma in this population^13^.

This study’s objective was to determine the dimensionality and internal consistency of the tuberculosis stigma instrument in patients living with TB in Santa Marta, Colombia.

## Method

A psychometric or validation study was carried out without a gold standard. This type of design uses statistical tests to evaluate the construct validity of an instrument, which allows us to approximate the reliability of the measurements.

### Participants

One hundred and twenty-two patients between the ages of 18 and 75 (M=40.3, SD=14.9) participated with complete primary or secondary schooling with reading and writing skills to complete the scales. The highest percentage of patients were men, single, living in low-income areas, pulmonary tuberculosis, and without HIV co-infection. More details of the participants in Table 1. This sample size was adequate for a validity and reliability analysis at the rate of ten participants for each item^20^.

**Table 1.**
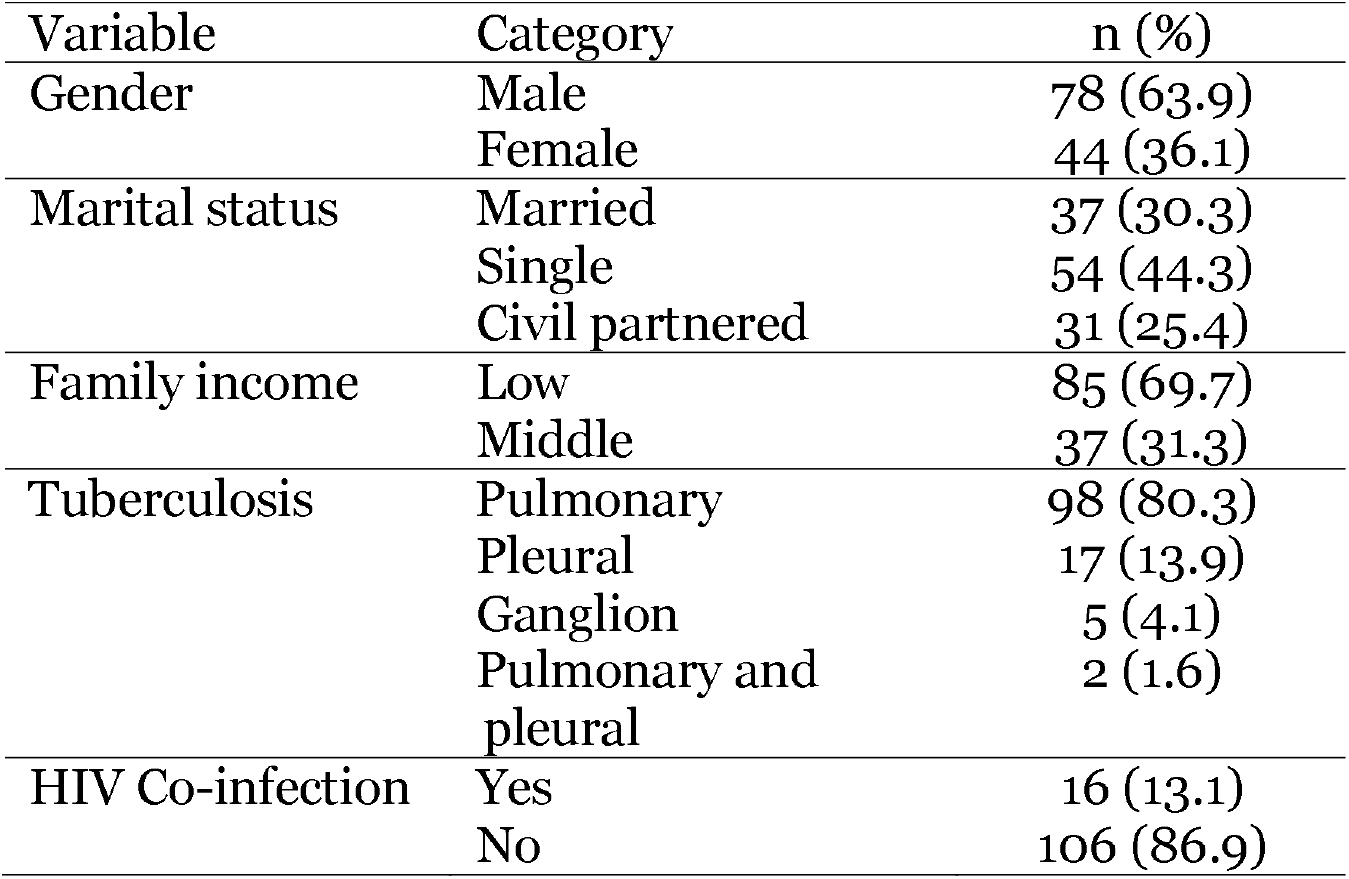
Description of participants (N = 122).

### Instruments

The Tuberculosis-Related Stigma Scale has two subscales, the first for perceived stigma and the second for internalized stigma. Each item offers four responses: Strongly disagree (score 1), disagree (score 2), agree (score 3), and strongly agree (score 4). The perceived stigma scale allows scores between 11 and 44 and the internalized stigma scale between 12 and 48. In both scales, the higher the score, the higher the level of stigma^16^.

### Procedure

The patients were invited to participate in the outpatient service. The study’s objectives, the anonymity of the response, the confidentiality of the data, and the utterly voluntary participation were explained. Participants self-filled out the questionnaires after brief instruction.

### Statistical analysis

The descriptive analysis of the scale included the observation for each item and the total scores of the mean (standard deviation) and the corrected correlation of the item with the total score. In the exploratory factor analysis, the Kaiser Meyer-Olkin’s sample adequacy test (KMO) and Bartlett’s sphericity test were observed, necessary to test whether a set of items underlies a construct. The extraction was carried out with the maximum likelihood method. Eigenvalues, explained variances, communalities, and loadings were observed.

In the confirmatory factor analysis, five indicators were calculated: Satorra-Bentler chi-square (X^2^/df), Root Mean Square Error of Approximation (RMSEA) and 90% confidence interval (90%CI), Comparative Fit Index (CFI), Tucker-Lewis index (TLI), and Standardized Mean Square Residual (SMSR). Chi-square with probability greater than 0.05 or X^2^ / df ratio less than five were expected,^21^ RMSEA and SMSR less than 0.08, and CFI and TLI values were more significant than 0.90. Dimensionality is accepted if three or more calculated coefficients are adequate^22^.

The coefficients of Cronbach’s alpha and McDonald’s omega were used to estimate internal consistency. McDonald’s omega is a better indicator of internal consistency when items show significant differences in factor loadings. Internal consistency is adequate if it shows values between 0.70 and 0.95^23^. These coefficients were calculated in the Jamovi program version 1.8.2^24^.

Student’s t was used to compare the scores in men and women. The Spearman correlation test was used to measure the nomological validity of the scales. A correlation between 0.30 and 0.60 would be an acceptable value^25^. The differential functioning of the item by gender was quantified with Kendall’s tau-b correlation. The performance is non-differential if the correlation is 0.20 and indicates that the response patterns are free of bias according to the reference variable^26^. It was calculated using STATA 13.0^27^.

### Ethical considerations

The project received approval from an institutional research ethics committee of a Colombian state university, according to Minutes 006 of the ordinary virtual session held on June 16, 2020. All the participants followed the guidelines and deontological norms enshrined in the Declaration of Helsinki^28^.

## Results

### Description of the items

Scores for the perceived stigma scale were observed between 11 and 44 (M=25.9, SD=6.2), the data distribution was skewed (V’=2.71, p=0.02). On the internalized stigma scale between 12 and 48 (M=28.7, SD=7.6), the distribution of the scores was symmetric (V’=1.85, p=0.11). The Spearman correlation between the scale scores was 0.50. The mean and standard deviation for each item are presented in Tables 2 and 3.

**Table 2.**
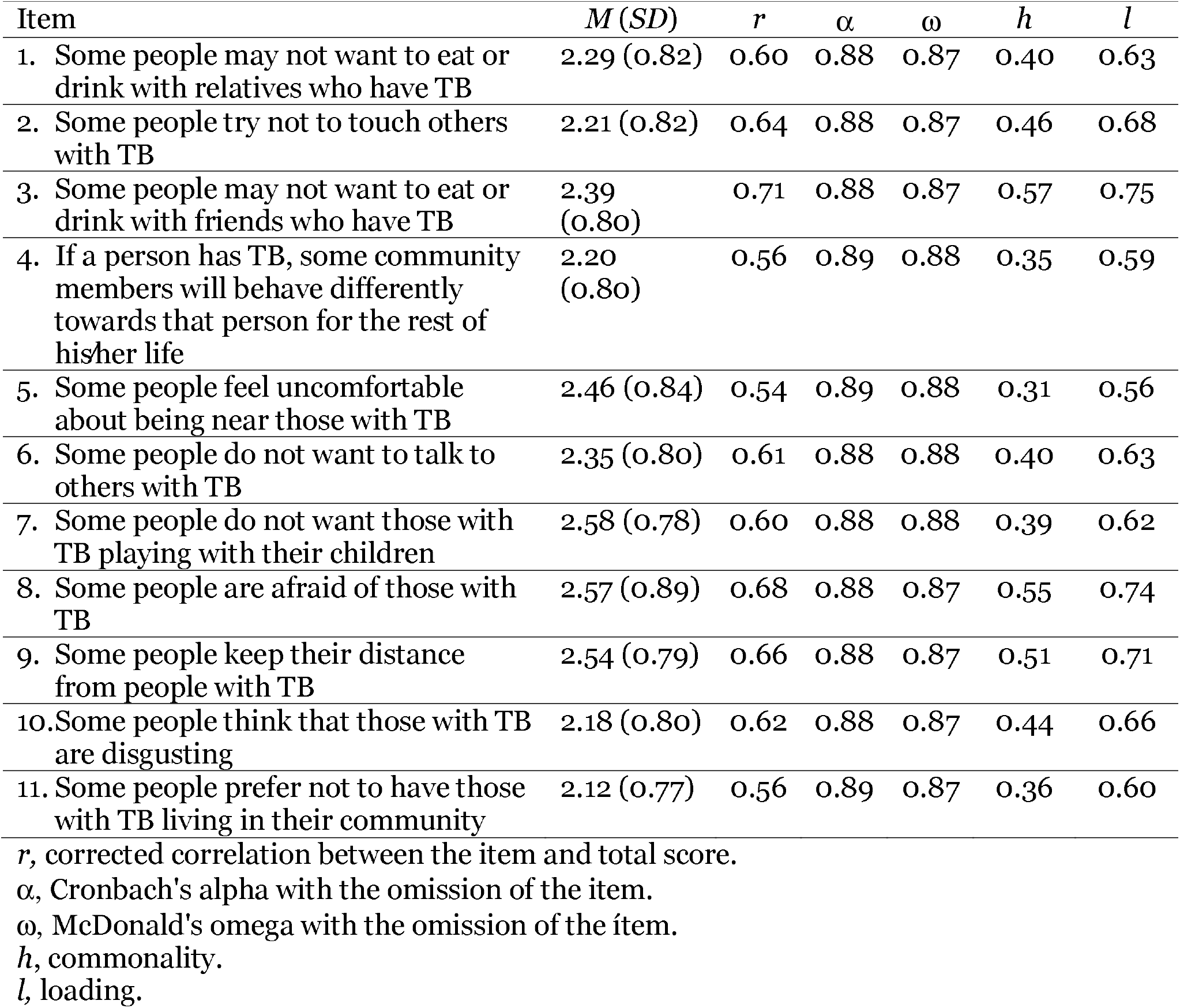
Descriptive of the perceived stigma scale items, corrected correlation, internal consistency if the item is omitted, communalities and loadings.

**Table 3.** Descriptive items of the internalized stigma scale.

| Item | *M* (*SD*) | *r* | $\alpha$ | $\omega$ | *h* | *l* |
| --- | --- | --- | --- | --- | --- | --- |
| 1. Some people who have TB are afraid to tell their families that they have TB | 2.36<br>(0.85) | 0.67 | 0.90 | 0.90 | 0.52 | 0.72 |
| 2. Some people who have TB are afraid of going to TB clinics because other people may see them there | 2.23<br>(0.76) | 0.66 | 0.90 | 0.90 | 0.51 | 0.71 |
| 3. Some people who have TB are afraid to tell others that they have TB because others may think that they also have AIDS | 2.52<br>(0.87) | 0.72 | 0.90 | 0.90 | 0.58 | 0.76 |
| 4. Some people who have TB feel guilty for getting TB because of their smoking, drinking, or other careless behaviours | 2.38<br>(0.96) | 0.65 | 0.90 | 0.90 | 0.48 | 0.69 |
| 5. Some people who have TB are worried about having AIDS | 2.26<br>(0.85) | 0.64 | 0.90 | 0.90 | 0.45 | 0.67 |
| 6. Some people who have TB feel guilty because their family has the burden of caring for them | 2.20<br>(0.90) | 0.71 | 0.90 | 0.90 | 0.57 | 0.75 |
| 7. Some people who have TB will choose carefully whom they tell about having TB | 2.56 (0.91) | 0.64 | 0.90 | 0.90 | 0.43 | 0.66 |
| 8. Some people who have TB lose friends when they share with them they have TB | 2.30<br>(0.88) | 0.68 | 0.90 | 0.90 | 0.50 | 0.71 |
| 9. Some people who have TB keep their distance from others to avoid spreading TB germs | 2.84<br>(0.93) | 0.57 | 0.91 | 0.91 | 0.34 | 0.58 |
| 10. Some people who have TB are afraid to tell those outside their family that they have TB | 2.48<br>(0.93) | 0.55 | 0.91 | 0.91 | 0.32 | 0.57 |
| 11. Some people who have TB feel alone | 2.20<br>(0.89) | 0.59 | 0.90 | 0.91 | 0.39 | 0.63 |
| 12. Some people who have TB feel hurt by how others react to knowing they have TB | 2.40<br>(0.94) | 0.65 | 0.90 | 0.90 | 0.45 | 0.67 |

*r*, corrected correlation between the item and total score.
$\alpha$ , Cronbach's alpha with the omission of the item.
$\omega$ , McDonald's omega with the omission of the item.
*h*, commonality.
*l*, loading.

### Exploratory and confirmatory factor analysis

The perceived stigma scale showed a KMO of 0.84 and Bartlett’s test of 644.7 (df=55, p<0.001). A single salient factor was retained with an eigenvalue of 5.29 that accounted for 48.1%. The internalized stigma scale showed a KMO of 0.87 and Bartlett’s test of 770.73 (df=66, p<0.001). A single salient factor with an eigenvalue of 6.07 responsible for 50.5% was retained. Communalities were observed above 0.30 and loadings above 0.56. These values are detailed in Tables 2 and 3. The confirmatory factor analysis of the sub-scales of the Tuberculosis Related Stigma Scale showed two acceptable goodness-of-fit indicators. See table 4.

**Table 4.**
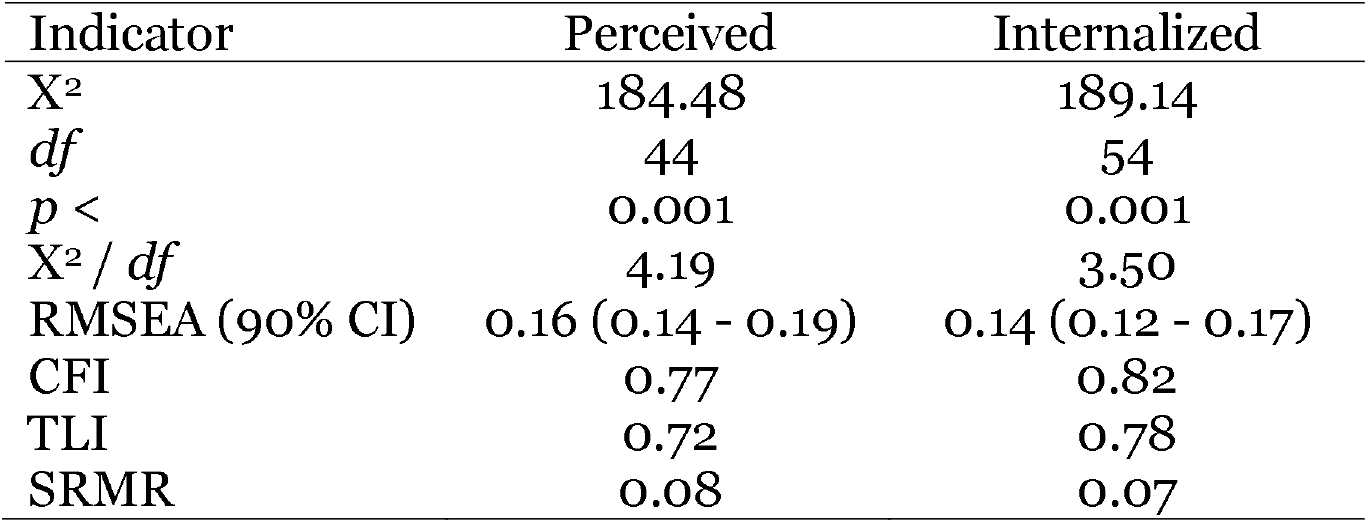
Goodness-of-fit indicators for the sub-scales.

### Internal consistency

The perceived stigma scale showed high consistency, Cronbach’s alpha, and McDonald’s omega of 0.89. The internalized stigma scale presented excellent consistency, Cronbach’s alpha and McDonald’s omega of 0.91. For both scales, the alpha and omega values were similar even after eliminating the item. See Tables 2 and 3.

### Gender response pattern

The perceived stigma scores were similar in men and women [M=25.3 (SD=6.1) versus M=25.1 (SD=6.28)]; Levene’s test for homogeneity of variances, F=0.11, p<0.74, t=1.05, df=120, p<0.30. Likewise, the results were comparable by gender on the internalized stigma scale [M=29.0 (SD=7.8) versus M=28.2 (SD=7.2)]; Levene’s test for homogeneity of variances, F=0.21, p<0.65, t=0.54, df=120, p<0.59 Differential item functioning was negative, Kendall’s tau B values were observed between 0.00 and 0.15. See Tables 5 and 6.

**Table 5.** Gender differential item functioning for the scale of perceived stigma.

| Item | $\tau$ |
| --- | --- |
| 1. Some people may not want to eat or drink with relatives who have TB | 0.11 |
| 2. Some people try not to touch others with TB | 0.07 |
| 3. Some people may not want to eat or drink with friends who have TB | 0.05 |
| 4. If a person has TB, some community members will behave differently towards that person for the rest of his/her life | 0.08 |
| 5. Some people feel uncomfortable about being near those with TB | 0.10 |
| 6. Some people do not want to talk to others with TB | 0.09 |
| 7. Some people do not want those with TB playing with their children | 0.10 |
| 8. Some people are afraid of those with TB | 0.08 |
| 9. Some people keep their distance from people with TB | 0.08 |
| 10. Some people think that those with TB are disgusting | 0.03 |
| 11. Some people prefer not to have those with TB living in their community | 0.02 |

**Table 6.** Gender differential item functioning for the scale of internalized stigma.

| Item | $\tau$ |
| --- | --- |
| 1. Some people who have TB are afraid to tell their families that they have TB | 0.01 |
| 2. Some people who have TB are afraid of going to TB clinics because other people may see them there | 0.02 |
| 3. Some people who have TB are afraid to tell others that they have TB because others may think that they also have AIDS | 0.00 |
| 4. Some people who have TB feel guilty for getting TB because of their smoking, drinking, or other careless behaviours | 0.15 |
| 5. Some people who have TB are worried about having AIDS | 0.05 |
| 6. Some people who have TB feel guilty because their family has the burden of caring for them | 0.08 |
| 7. Some people who have TB will choose carefully whom they tell about having TB | 0.05 |
| 8. Some people who have TB lose friends when they share with them they have TB | 0.03 |
| 9. Some people who have TB keep their distance from others to avoid spreading TB germs | 0.05 |
| 10. Some people who have TB are afraid to tell those outside their family that they have TB | 0.03 |
| 11. Some people who have TB feel alone | 0.01 |
| 12. Some people who have TB feel hurt by how others react to knowing they have TB | 0.05 |

## Discussion

In the present study, the Tuberculosis-Related Stigma Scale presented high internal consistency; however, the data did not adjust to the dimensionality proposed for the perceived and internalized stigma subscales.

These findings are similar to those reported in the few investigations with patients living with tuberculosis regarding internal consistency. In Brazil, Crispim et al.^17^ found Cronbach’s alpha of 0.70 for the perceived stigma subscale and 0.71 for the internalized stigma subscale in a sample of 83 participants. In Taiwan, Lee et al.^29^ found alpha values of 0.98 for the perceived stigma subscale and 0.95 for the internalized stigma subscale in 84 participants. In Mexico, Moya et al.^18^ reported Cronbach’s alpha of 0.91 on the perceived stigma subscale and 0.87 on the internalized stigma subscale. Previous research omitted information on McDonald’s omega. Currently, it is recommended to report at least two measures of reliability. McDonald’s omega is interpreted the same as Cronbach’s alpha and is a more reliable statistic of the internal consistency of a scale when the principle of tau equivalence assumed for the calculation of Cronbach’s alpha is infringed^23^.

Concerning dimensionality, the data did not fit the one-dimensional sub-scale of perceived stigma or internalized stigma in the present investigation. These data are consistent with that reported by Van Rie et al.^16^; the researchers found a questionable two-dimensional structure since it showed one of three indicators within the desirable value (TLI of 0.94, TLI of 0.88, and RMSEA of 0.11). These findings show the need to refine the Tuberculosis-Related Stigma Scale until reaching indicators of goodness of fit that corroborate the theoretical unidimensionality^22^.

### Practical implications

Valid and reliable instruments are essential for the measurement of TB-SDC. Historically, some diseases have been the target of SDC that can be configured as a significant stressor in affected people’s lives^9,10,30^. Frequently, the effect of SDC is much more devastating than the diagnosis of the disease^8^. TB-SDC delays the consultation due to symptoms suggestive of TB and, likewise, it is associated with less adherence to the drug treatment plan^31,32^. In general, TB-SDC worsens the clinical prognosis of the disease^31^. Furthermore, SDC increases the risk of distress and psychological suffering, which leads to a higher risk of depression and suicidal behaviours.^15,33^ Consequently, all health personnel who care for people who live with TB, doctors, nurses, psychologists, social workers, and health promoters should actively explore TB-SDC. They should initiate or refer to other professionals specialized in psychological interventions to reduce the negative impact of TB-SDC^34^.

### Strengths and limitations of the study

This study is the first study to show the psychometric evaluation of the Tuberculosis-Related Stigma Scale in a sample of Colombian patients. However, it has the limitation that a refinement process can be carried out due to the limited sample size. In these cases, the result is more reliable if there is a sample of 400 or more participants^20^. Likewise, it is necessary to explore the performance of the Tuberculosis-Related Stigma Scale in other samples since the scales can be highly sensitive to the sociocultural characteristics of the participants^35^.

### Conclusions

It is concluded that the subscales of the Tuberculosis-Related Stigma Scale have high internal consistency but poor indicators of one-dimensionality. Therefore, it is recommended that future research carry out a refinement process with a large sample of participants and retain better-performing items.

## Data Availability

All data produced in the present study are available upon reasonable request to the authors.

## Acknowledgement of conflicts of interest

The authors have no conflicts of interest to declare.

## References

1. World Health Organization. Global tuberculosis report 2020. Geneva: World Health Organization; 2020. (Acceso: Marzo 30, 2021). https://apps.who.int/iris/bitstream/handle/10665/336069/9789240013131-eng.pdf

2. Stop TB Partnership. The Global Plan to End TB 2018-2022.Internet Geneva: Stop TB org; 2021. (Acceso: Marzo 30, 2021). http://www.stoptb.org/assets/documents/global/plan/GPR_2018-2022_Digital.pdf.

3. Ministerio de Salud y Protección Social República de Colombia y la Organización Panamericana de la Salud. Strategic Plan “Towards the end of Tuberculosis” Colombia 2016-2025. Adaptation tools of the Post-2015 Tuberculosis Free Colombia Strategic Plan. Agreement 519 of 2015 Colombia. Bogotá: 2016.

4. TB Report App. Geneva: World Health Organization; 2020.

5. Craig GM, Daftary A, Engel N, O’Driscoll S, Loannaki A. Tuberculosis stigma as a social determinant of health: a systematic mapping review of research in low incidence countries. IJID. 2017;2017:90–100.

6. Restrepo-Zea JH, Silva-Maya C, Andrade-Rivas F, VH-Dover, R. Access to health services: Analysis of barriers and strategies in Medellín, Colombia. Rev Gerenc Polit Salud. 2014;13(27):242–265.

7. Mukerji R, Turan JM. Exploring Manifestations of TB-Related Stigma experienced by women in Kolkata, India. Global Health. 2018;84(4):727–735.

8. Campo-Arias A, Herazo E. Stigma, prejudice, and discrimination in mental health Rev Cienc Biomed. 2013;4(1):9–10.

9. Pescosolido BA, Martin JK. The stigma complex. An Rev Soc. 2015;2015:87–116.

10. Tyler IE, Slater T. Rethinking the sociology of stigma. Sociol Rev. 2018;66(4):721–743.

11. Coreil J, Mayard G, Simpson KM, Lauzardo M, Zhu Y, Weiss M. Structural forces and the production of TB-related stigma among Haitians in two contexts. Soc Sci Med. 2010;71(8):1409–1417.

12. Duko B, Bedaso A, Ayano G, Yohannis Z. Perceived stigma and associated factors among patient with tuberculosis, Wolaita Sodo, Ethiopia: Cross-sectional study. Tuberc Res Treat. 2019:5917537.

13. Courtright A, Norris A. Tuberculosis and stigmatization: pathways and interventions. Public Health Rep. 2010;125(4):34–42.

14. Yin X, Yan S, Tong Y, Xin P, Yang T, Lu Z, et al. Status of tuberculosis-related stigma and associated factors: a cross-sectional study in central China. TMIH. 2018;23(2):99–205.

15. Koyanagi A, Vancampfort D, Carvalho AF, DeVylder J, Pizzol D, Veronese N, et al. Depression comorbid with tuberculosis and its impact on health status: cross-sectional analysis of community-based data from 48 low- and middle-income countries. BMC Medicine. 2017;2017:209.

16. Van Rie A., Sengupta S, Pungrassami P, Balthip Q, Choonuan S, Kasetjaroen Y, et al. Measuring stigma associated with tuberculosis and HIV/AIDS in southern Thailand: exploratory and confirmatory factor analyses of two new scales. Trop Med Int Health. 2008;2008:21–30.

17. Crispim J, Mosna M, Yamamura M, Paschoal M, Garcia M, Dos Santos C, et al. Cultural adaptation of the Tuberculosis-related stigma scale to Brazil. Cienc Saúde Colet. 2016;21(7):2233–2242.

18. Moya EM, Biswas A, Chávez Baray SM, Martínez O, Lomeli B. Assessment of stigma associated with tuberculosis in Mexico. Public Health Action. 2014;2014:226–232.

19. Upegui LD, Orozco LC.. Design of an instrument to measure stigma towards tuberculosis Salud UIS. 2014;2014:22–34.

20. Kahn JH. Factor analysis in counselling psychology, research, training, and practice: Principles, advances, and application. Counsel Psychol. 2006;34(5):684–718.

21. Wheaton B, Muthen B, Alwin DF, Summers GF. Assessing reliability and stability in panel models. In: Heise DE. Sociological methodology. San Francisco: Jossey-Bass, 1977. pp. 84–136.

22. Hu LT, Bentler PM. Cutoff criteria for fit indexes in covariance structure analysis: Conventional criteria versus new alternatives. Struct Equat Model. 1999;6(1):1–55.

23. Keszei AP, Novak M, Streiner DL. Introduction to health measurement scales. J Psychosom Res. 2010;68(4):319–323.

24. Jamovi Project. Jamovi (Version 1.8.2), 2021. Retrieved from https://www.jamovi.org

25. Gupta B. Interview questions in business analytics. Berkeley: Apress 2016; pp. 45–55.

26. Hambleton RK. Good practices for identifying differential item functioning. Med Care. 2006;44(11):S182–S188.

27. StataCorp. Stata Statistical Software: Release 14. College Station, TX: StataCorp LP, 2015.

28. World Medical Association. WMA Declaration of Helsinki – Ethical principles for medical research involving human subjects. Seoul, Korea: WMA, 2018.

29. Lee L-Y, Tung H-H, Chen S-C, Fu C-H. Perceived stigma and depression in initially diagnosed pulmonary tuberculosis patients. J Clin Nurs. 2017;26(23–24):4813–4821.

30. Ascuntar JM, Gaviria MB, Uribe L, Ochoa J. Fear, infection and compassion: Social representations of tuberculosis in Medellin, Colombia, 2007. Int J Tuberc Lung Dis. 2010; 14(10):1323–1329.

31. Juniarti N, Evans D. A qualitative review: the stigma of tuberculosis. J Clin Nurs. 2011;20(13 14):1961–1970.

32. Chakrabartty A, Basu P, Ali KM, Ghosh D. Tuberculosis related stigma attached to the adherence of Directly observed treatment short course (DOTS) in West Bengal, India. Indian J Tuberc. 2019;66(2):259–265.

33. Peltzer K, Louw J. Prevalence of suicidal behaviour & associated factors among tuberculosis patients in public primary care in South Africa. Indian J Med Res. 2013;138(2):194–200.

34. Weiss MG, Ramakrishna J, Somma D. Health-related stigma: rethinking concepts and interventions. Psychol Health Med. 2006;11(3):277–287.

35. Carvajal A, Centeno C, Watson R, Martínez M, Sanz Á. How is an instrument for measuring health to be validated? An Sist Sanit Navar. 2011;34(1):63–72.

